# BDNF, inflammatory and oxidative levels in treatment-naïve ADHD children treated with methylphenidate: An open cohort protocol

**DOI:** 10.1101/2023.03.03.23286780

**Authors:** Marina Silva de Lucca, Laira Lopes Tonon, Jordânia Alves Ferreira, Bárbara Silva Cabral, Cleuberton Kenedy Oliveira Raimundo, Sílvia Almeida Cardoso, Débora Marques de Miranda

## Abstract

The attention-deficit hyperactivity disorder (ADHD) has a complex etiology, involving the interaction between biological, genetic, and environmental factors. The ADHD pathophysiology remains unknown even though there are hypotheses that inflammatory, hormonal, oxidative and neurotrophic factors are associated. This clinical trial aims to evaluate the contribution of brain derived neurotrophic factor (BDNF), inflammatory and oxidative levels before and after 12 and 24 weeks of methylphenidate use.

**Methods:** Patients will be screened upon their entry into Child and Adolescent Psychiatry Teaching Outpatient Clinic of the Medical Course at the Federal University of Viçosa in Minas Gerais, Brazil. One hundred and fifty ADHD treatment-naïve children of both sexes, between 6–14 years old, will be invited to participate, after the ADHD diagnosis by an experienced psychiatrist and the child fulfilling the inclusion criteria. Children and their caregivers will answer questionnaires regarding mental health and the children will undergo neuropsychological tests, physical, nutritional and activity assessment, in addition to blood sampling at baseline, 12 and 24 weeks of methylphenidate use respectively.

**Discussion:** This clinical trial intends to verify how the pharmacological treatment changes the plasma BDNF, inflammatory and oxidative levels in treatment-naïve Brazilian children diagnosed for ADHD.

**Trial Registration:** Submitted for registration on Brazilian Registry of Clinical Trials (ReBEC). Trial identifier: 13612

Registry name: Níveis de neurotrofina, perfil inflamatório e oxidativo em crianças com TDAH tratadas com metilfenidato.

## Administrative information – Spirit 2013 Checklist

### Title {1}

BDNF, inflammatory and oxidative levels in treatment-naïve ADHD children treated with methylphenidate: An open cohort protocol.

### Trial registration

#### Registry {2a}

Submitted for registration on Brazilian Registry of Clinical Trials (ReBEC). Trial identifier: 13612.

Registry name: Neurotrophin level, inflammatory and oxidative profile in ADHD children treated with methylphenidate.

#### Data Set {2b}

The Universal Trial Number (UTN) is U1111-1285-2908.

Date recruitment began: 23/10/2020; Approximate date when recruitment will be completed: 29/11/2022.

### Protocol version {3}

First version.

Submitted for registration on Brazilian Registry of Clinical Trials (ReBEC). Trial identifier: 13612.

### Funding {4}

National Council for Scientific and Technological Development (CNPQ).

Research Support Foundation of the State of Minas Gerais (FAPEMIG - APQ-01023/18)

### Roles and Responsibilities {5}

#### Contributorship {5a}

Marina Silva de Lucca. Master in Physical Education and PhD student in Health Sciences, Faculty of Medicine, UFMG.

Laira Lopes Tonon. Medicine student (UFV). Research volunteer member. Bárbara Silva Cabral. Medicine student (UFV). Research volunteer member.

Jordânia Alves Ferreira. Medicine student (UFV). Scientific initiation scholarship Cleuberton Kenedy Oliveira Raimundo. Medicine student (UFV). Research volunteer member.

Silvia Almeida Cardoso. Master and Doctor in Immunology (USP). Associate Professor of Nursing and Medicine, Faculty of Medicine, UFV.

Débora Marques de Miranda. Master and Doctor in Biochemical and Molecular Pharmacology (UFMG). Associate Professor of Pediatrics, Faculty of Medicine, UFMG. ML is the Chief Investigator; she conceived the study, led the proposal and protocol development. LLT, BSC, JAF and CKOR contributed to development of the proposal and methodology. SC e DM contributed to study design and to development of the proposal and methodology. All authors read and approved the final manuscript.

#### Sponsor contact information {5b}

Not applicable.

#### Sponsor and Funder {5c}

This funding source had no role in the design of this study and will not have any role during its execution, analyses, interpretation of the data, or decision to submit results.

#### Committees {5d}

##### Principal Investigator and Research Physician

Design and conduct the clinical appointments: Marina Silva de Lucca.

Preparation of protocol and revisions: Marina Silva de Lucca, Sílvia Almeida Cardoso, and Débora Marques de Miranda.

Organizing steering committee meetings: Marina Silva de Lucca and Débora Marques de Miranda.

Publication of study reports: Marina Silva de Lucca, Sílvia Almeida Cardoso, and Débora Marques de Miranda.

##### Steering committee

The composition of the trial committee includes all authors.

Agreement of final protocol: Marina Silva de Lucca, Sílvia Almeida Cardoso, and Débora Marques de Miranda.

All lead investigators will be steering committee members.

Recruitment of patients: Laira Lopes Tonon, Bárbara Silva Cabral and Jordânia Alves Ferreira, Cleuberton Kenedy Oliveira Raimundo.

Reviewing progress of study and if necessary, agreeing changes to the protocol and/or investigators brochure to facilitate the smooth running of the study: Marina Silva de Lucca and Débora Marques de Miranda.

##### Trial Management Committee (TMC)

Study planning; Organization of steering committee meetings: Marina Silva de Lucca and Débora Marques de Miranda.

Data verification: Marina Silva de Lucca.

Endpoint adjudication: Marina Silva de Lucca and Débora Marques de Miranda.

##### Data Management team

Data entry, data verification: Laira Lopes Tonon, Bárbara Silva Cabral, Jordânia Alves Ferreira, Cleuberton Kenedy Oliveira Raimundo and Marina Silva de Lucca.

##### Lead Investigators

Marina Silva de Lucca, Sílvia Almeida Cardoso, and Débora Marques de Miranda.

## Schedule of enrolment, interventions, and assessments

### Introduction

#### Background and rationale {6a}

Attention deficit hyperactivity disorder (ADHD) is a neurodevelopmental disorder characterized by symptoms of inattention, hyperactivity, and impulsivity that appear in childhood and usually persist into adulthood, causing some degree of dysfunction in daily activities [1,2,3]. The heterogeneous clinical presentations of ADHD are a hallmark of this disorder [4] important negative outcomes are reported, both in terms of morbidity and mortality [3].

ADHD has a complex etiology, involving the interaction between genetic, biological and environmental factors. However, its pathophysiology remains unknown [5]. There are hypotheses that inflammatory, oxidative, hormonal, and neurotrophic factors are associated, and they have mutual interactions. [3, 6].

Different ADHD risk factors, such as prematurity, paternal smoking, exposure to pesticides and lead may be associated with common pathophysiological pathways, such as inflammation, oxidative stress, and neurotrophic factors. Likewise, intrauterine pro-inflammatory factors during the gestational period may be associated with restricted intrauterine growth, miscarriage, premature birth, placenta abruption, neurological damage, some of which are risk factors for ADHD [7,8,9, 10, 11, 12]. Neuroinflammation in physiological and pathological conditions may activate microglia, astrocytes, oligodendrocytes, and ependymal cells. In addition, it may increase proteases, glutamate, reactive oxygen species, nitric oxide, chemokines, toxic cytokines, prostaglandins, and may induces infiltration of T and B neutrophils, monocytes/ macrophages and dendritic cells. When activated, these cells release pro-inflammatory cytokines that increase neuroinflammation [13]. Some cytokines are elevated in ADHD, some of which are related to the severity of symptoms [13, 14, 15]. A systematic review on the level of cytokines in peripheral blood showed increased levels of cytokines-interleukin-6 (IL-6) in patients with ADHD, especially in those younger than eighteen years and not on medication, when compared to a healthy control group [13, 16]. However, another systematic review found no difference in IL-6 levels when comparing children with and without ADHD [17]. Furthermore, patients with ADHD, especially in those younger than eighteen years and not on medication, had lower levels of tumor necrosis factor-alpha human (TNF-alpha) compared to healthy controls [16, 17]. The results regarding cytokines-interleukin-10 (IL-10) were contradictory and may be increased or unchanged in children with ADHD compared to children without the pathology [13, 16, 17]. ADHD treated patients seem to have a decrease in interferon gamma (IFN-gamma) and interleukin-13 (IL-13) levels, compared to those naïve to treatment [13].

Therefore, neuroinflammation could alter the blood-brain barrier, neurotransmitter metabolism, increase oxidative stress, and neurodegeneration [14, 15]. Animals and humans with ADHD show, in different brain structures, an increase in reactive oxygen species without an adequate antioxidant response, resulting in an increase in oxidative stress. This negative imbalance in ADHD may be explained in the central nervous system due to the large consumption of oxygen by neurons associated with an antioxidant defense system of modest action and a constitution rich in lipids. As a result, the brain has difficulty regulating excess reactive oxygen species, making it more susceptible to damage [15, 18, 19, 20, 21, 22]. Therefore, there is evidence of increased oxidative stress in ADHD patients [19, 20, 21 22, 23, 24].

ADHD may be associated with a decrease in ascorbate, catalase, albumin and bilirubin and an increase in uric acid. So, it may contribute to oxidative stress increase [25]. On the other hand, treatment with methylphenidate seems to increase catalase levels. Other study demonstrated that the activity of superoxide dismutase (SOD), an important antioxidant defense enzyme, is lower in patients with ADHD [25].

Increased oxidative stress may be associated with reduced levels of BDNF, as well as reduced oxidative stress with increased BDNF [26]. Therefore, changes in the BDNF expression at critical moments during development may promote a cascade of events interfering with the brain maturation of some regions, being a substrate for altered response to stress in adulthood and development of neuropsychiatric disorders [26]. Another evidence of intense oxidative stress is the modulation of telomere size and shortening during life [27] and telomere shortening is present in ADHD children [28]. So, telomere length may be a potential biomarker of the ADHD symptoms burden in families affected by this neurodevelopmental disorder [29].

Studies with rats have been trying to observe the results of chronic use of methylphenidate in brain cells [30, 31, 32]. Chronic use of methylphenidate in healthy rats caused oxidative damage in the brain of young rats [30]. Furthermore, the chronic use of methylphenidate by adult rats induced oxidative stress and inflammation in the hippocampus, causing cellular damage in this area [31]. Increased oxidative stress was also observed in spontaneously hypertensive adult rats (animal version of ADHD) [32].

In ADHD children and adolescents’ study, low levels of nitric oxide were found in baseline, which did not change significantly with the use of methylphenidate over ten weeks [33]. Guney et al. (2015) [33] found that methylphenidate may reduce oxidant levels and increase antioxidant levels in children and adolescents.

Studies have evaluated BDNF levels in individuals with ADHD, comparing them with typical children [17, 35, 36, 37, 38, 39, 40, 41, 42, 43, 44, 45, 46, 47, 48, 49, 50, 51] and after stimulant treatment [40, 45, 50, 51, 52], with divergent results.

Due to the controversial studies and the importance of treating ADHD, this protocol study aims to explain the methodology that will evaluate BDNF, inflammatory and oxidative levels, their associated factors in ADHD treated children for 6 months with methylphenidate.

#### Choice of comparators {6b}

BDNF, inflammatory markers and oxidative stress levels of the ADHD children will be compared before and after methylphenidate use. Therefore, the same group of children will be compared in three different times: before starting methylphenidate and after 12 and 24 weeks of methylphenidate use. All patients will be in follow-up at the child and adolescent psychiatry outpatient clinic, receiving pharmacological treatment for ADHD. Local health system provides free methylphenidate (generic medication) in the presentation of 10mg, immediate-release tablets.

Control groups (ADHD children without methylphenidate use) will not be possible, because it would be unethical to deprive them of effective treatment, capable of reducing not only morbidity, but also mortality.

### Objectives {7}

The objectives of this open cohort clinical trial will be:

1. Evaluate the BDNF, inflammatory and oxidative levels before and after 12 and 24 weeks of methylphenidate use in treatment-naïve ADHD children.
2. Investigate if there is a moderating effect of the sociodemographic data, ADHD presentation, comorbidity presence, caregiver psychopathology, parenting styles, emotional regulation level on BDNF, cytokines and oxidative stress level, telomere length, and other variables from the survey instruments detailed throughout this protocol.

### Trial Design {8}

This trial is designed as an open cohort, single center, with convenience sample.

## Methods: Participants, Interventions, Outcomes

### Study setting {9}

All subjects will be invited to the child psychiatry teaching outpatient clinic at the Federal University of Viçosa (UFV), in the state of Minas Gerais, Brazil.

### Eligibility criteria {10}

### Participants inclusion criteria: {10}

Treatment-naïve ADHD children, of both sex in outpatient treatment; age between complete 6 and incomplete 15 years old. Children must have criteria for ADHD diagnosis according to the DSM-5 and must not have chronic diseases or use of any other medications that may interfere with the immune system or BDNF. Children who need iron or vitamin replacement may be included. The following comorbidities may be included: oppositional defiant disorder (ODD), tic disorder, enuresis, encopresis, skin picking disorder. Autism spectrum disorder (ASD) level 1, Classification of Diseases and Related Health Problems (ICD) 10 F84.5, could be included if the diagnosis occurs during the study. All parents must sign the free and informed consent form, just like children should also sign the assent form. The terms will be delivered during the initial assessment of the participants, if they wish to participate voluntarily in the research.

### Participants exclusion criteria: {10}

Families and children that refusal to participate in the research or refusal to use stimulant medications; presence of autoimmune, neurodegenerative diseases, and immunodeficiencies; intellectual disability, severe clinical or psychiatric comorbidities, except autism spectrum disorder level 1 (ICD 10 F84.5), ODD, tics, trichotillomania, skin picking disorder, enuresis, and encopresis; previous or current use of stimulants or other psychiatry medications; current use of medications for chronic diseases or that interfere in immune system or BDNF levels (antidepressants for example); girls who have menstruated; stimulant contraindication use.

### Participant timeline: {13, 14, 15, 18}

Participants will be included from October 2020 to November 2022, targeting a sample size of 150 children, based on sample calculation with analysis power between 80% and 95% of accuracy. This sample calculation was based on an article of Akay et al., 2017 [37].

They will be forwarded by physicians, schools and psychologists from the city and health region. To reach the sample number, the health units, as well as the municipal education department were communicated about the research and how the children could be referred for screening. There was also publicity on the local radio and the website of the Federal University of Viçosa.

Children who are eligible for the study will be categorized by ADHD presentation, disorder severity, with or without Oppositional Defiant Disorder (ODD), with or without autism spectrum disorder (ASD), at any emotional dysregulation level. Their caregivers will be sorted by parenting style.

Overall, each participant will make 8 to 10 visits during the study period. The research procedure for participants at each visit are described in Fig. 2.

**Fig 1.**
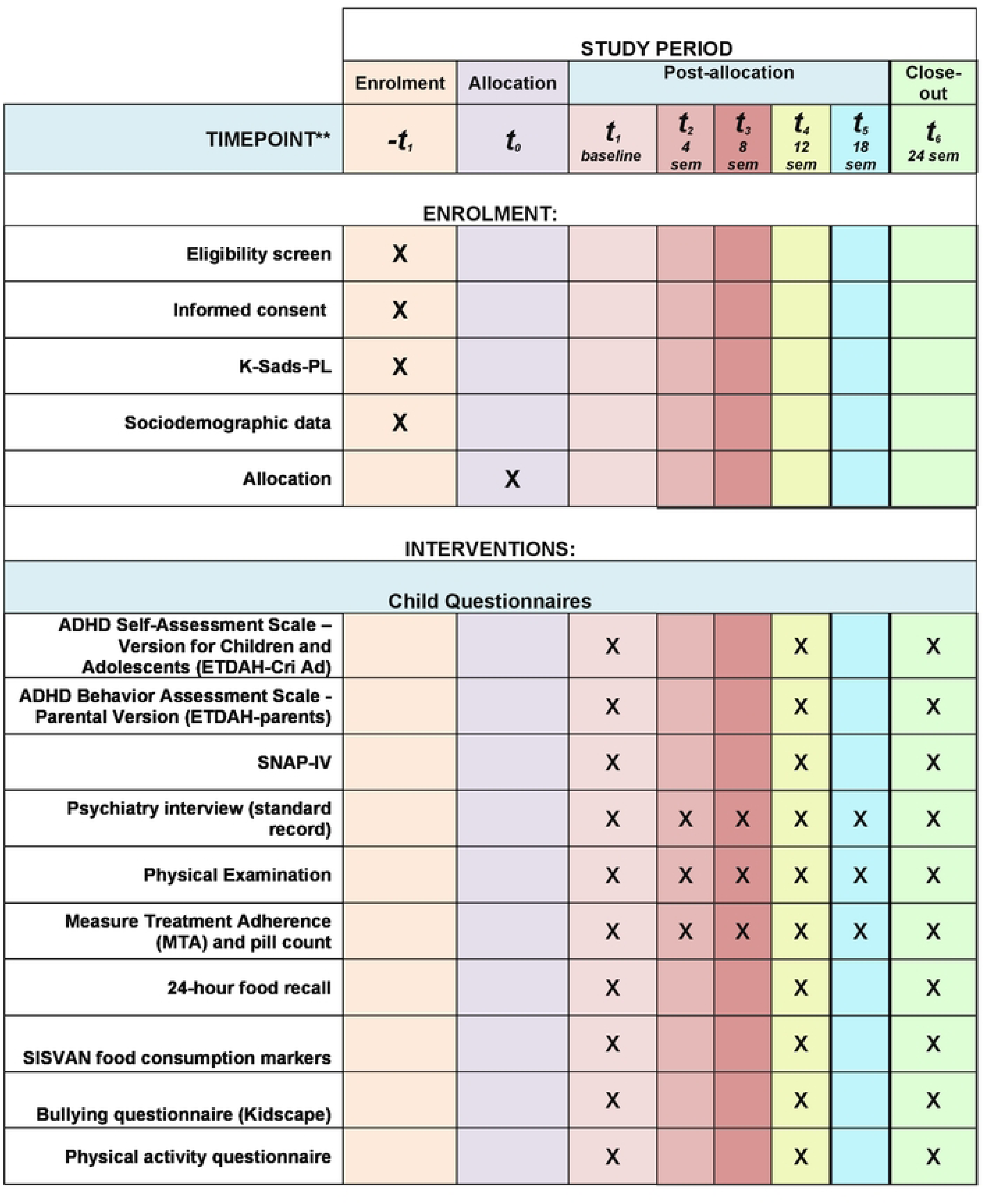
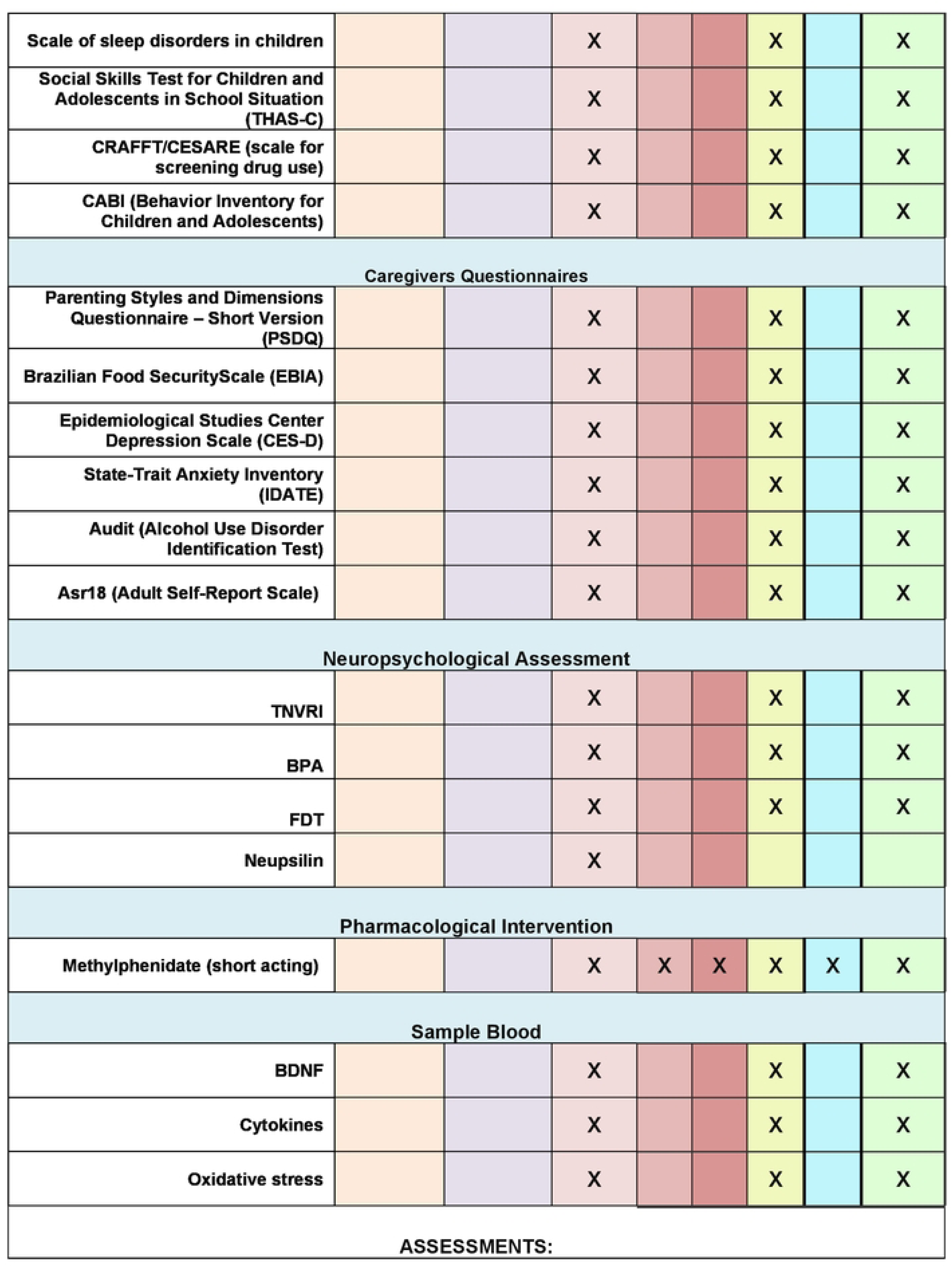
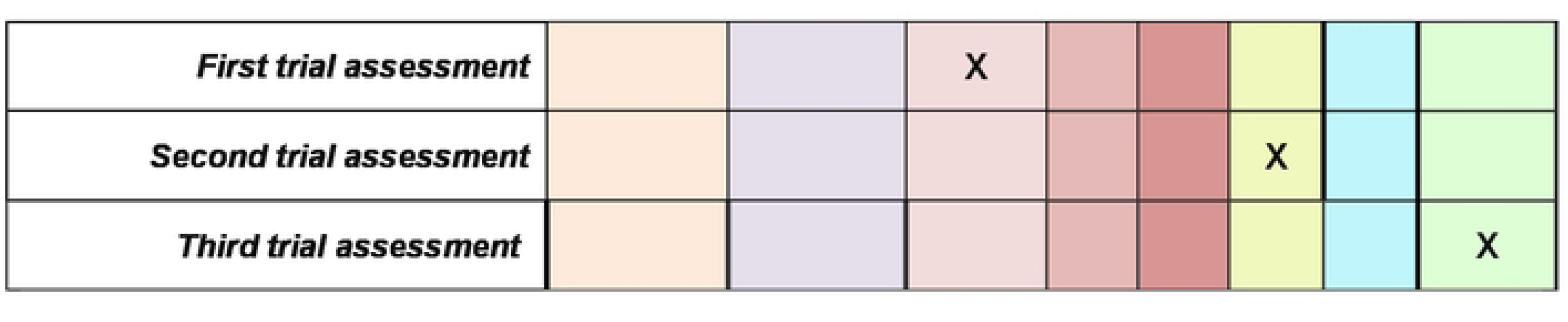
Schedule of enrolment, interventions, and assessments.

**Fig. 2.**
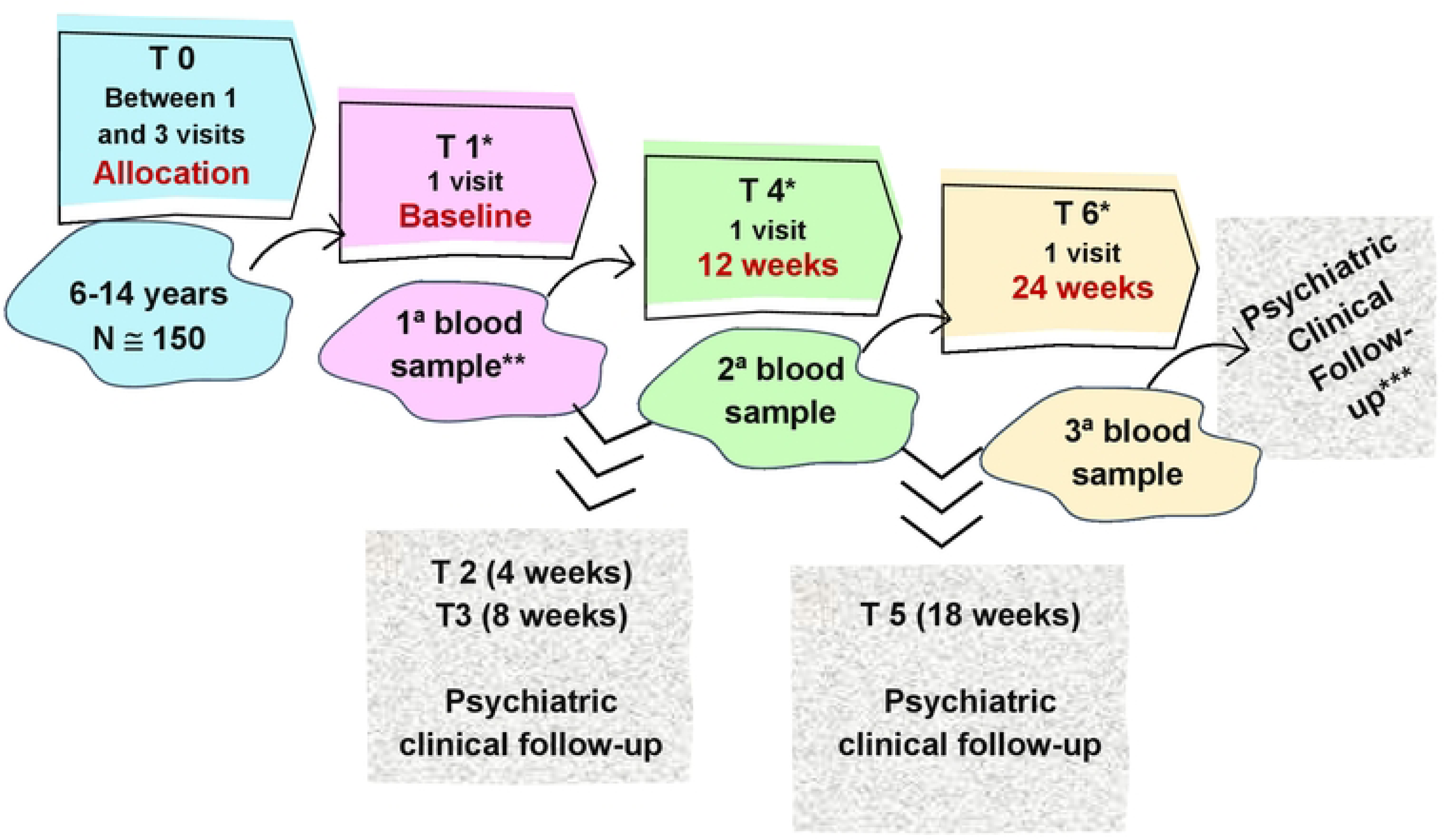
The participants visit. *Assessment points (Psychiatric clinical follow-up, Nutrition assessment, Neuropsychological assessment, Sample Blood, Questionnaires and Scales) **Methylphenidate prescription after first blood sample. ***Children who finished the trial times will be transferred to the general outpatient clinic of child psychiatry at the UFV medical school.

The initial assessment includes the semi-structured interview Kiddie-Schedule for Affective Disorders and Schizophrenia – Present and Lifetime Version (K-Sads-PL 2013) [53].

To assess the severity of ADHD symptoms, the SNAP-IV scale, Swanson, Nolan e Pelham version [54], and the Assessment Scale of Childhood and Adolescent Behaviors in ADHD in a Family Environment (ETDAH-parents) [55] and ADHD Self-Assessment Scale-Version for Children and Adolescents (ETDAH-Criad) [56] were answered by parents and children. The SNAP-IV scale has 26 items, which also investigates symptoms of a common comorbidity that is Defiant and Oppositional Disorder. [54]. The ETDAH scale has one version for parents and another for the children. [55, 56]. The parent subscale has four subscales that investigates symptoms of inattention, hyperactivity/impulsivity, emotional regulation, and adaptive behavior. The children subscale has 2 subscales that investigates inattention and hyperactivity/impulsivity symptoms.

The diagnosis will be confirmed by a interview done by a Child and Adolescent Psychiatrist. Sociodemographic data was obtained from the Brazil Economic Ranking Criterion and “Background information” part of the K-Sads-PL instrument (K-Sads-PL 2013) [53, 57, 58]. A standard record for the clinical history was constructed with data on pregnancy, childbirth, neuropsychomotor development, and the child’s previous and family history.

For the care of children at the outpatient clinic, an anamnesis model was built. This anamnesis also includes a habitual physical activity questionnaire [59], Brazilian food safety scale [60], SISVAN food consumption markers questionnaire [61], food frequency questionnaire [62, 63], bullying questionnaire (Kidscape) [64], measurement questionnaire adherence to treatment [65], CRAFFT/CESARE instrument (Car; Relax; Alone; Forget; Family/Friends; Trouble acronym) [66].

Situations of violence and/or health identified during the research will be duly forwarded to the necessary assistance network, such as justice, other medical specialties, social services, and others. The caregiver is screened for symptoms of depression, anxiety, ADHD, and alcohol use [67, 68, 69, 70]. In case of positive screening, evaluation by an adult psychiatrist will be offered.

### Interventions {11a, 18}

#### Screening visits

Medical students trained by an experienced child and adolescent psychiatrist and under her supervision, will apply in the first assessment (between one and three visits): Fig. 1. Schedule of enrolment, interventions, and assessments.

\# Multimodal Treatment Assessment Study – Swanson, Nolan e Pelham (MTA-SNAP-IV) scale, that evaluates symptoms of attention deficit/hyperactivity disorder and oppositional defiant disorder in children and adolescents [54]
\# Kiddie Schedule for Affective Disorders and Schizophrenia for School Aged Children – Lifetime Version (K-Sads-PL 2013) [53]
\# Sociodemographic questionnaire, including the Brazil Economic Ranking Criterion and the background information of the K-Sads-PL 2013 [53,57,58]
\# ADHD Behavior Assessment Scale – Parents Version (ETDAH-parents) [55]
\# Bullying questionnaire [64]
\# Physical activity questionnaire [59]
\# SISVAN food consumption markers [61]
\# Brazilian Food Security Scale (EBIA) [60]
\# Scale of Sleep disorders in children [72]
\# Behavior Inventory for Children and Adolescents (CABI) [73]
\# Parenting Styles and Dimensions Questionnaire – Short Version (PSDQ) [74]
\# Epidemiological Studies Center Depression Scale (CES-D) [67]
\# State-Trait Anxiety Inventory (IDATE) [68]
\# Adult Self-Report Scale (ASRS-18) [69]
\# Alcohol Use disorders identification test (AUDIT) [70]
\# Standardized psychiatric clinic interview with standard record.
\# Physical examination.

Psychology students, under supervision of an experienced psychologist in neuropsychology will apply:

\# Scale for screening drug use CRAFFT/CESARE [66]
\# Social Skills Test for Children and Adolescents in School Situation (THAS-C) [75]
\# ADHD Self-Assessment Scale – Version for Children and Adolescents (ETDAH-CriAd) [56]
\# Standardized and validated psychological tests for the Brazilian population.

- Non-Verbal test of Children’s Reasoning (TNVRI) [76]
- Battery for Attention Assessment (BPA) [77, 78]
- Five digits teste (FDT) [79]
- Brief Child Neuropsychological Assessment Instrument (NEUPSILIN-inf) [80]

Nutrition’s student under supervision of an experienced nutritionist will apply:

\# 24-hour food recall [81, 82]

### Assessment points {11a, 18a}

The assessment points occur in three timepoints: baseline (before methylphenidate initial use) and after 12 and 24 weeks of methylphenidate use. Fig 1 and 2.

In the first assessment point the blood sample is collected, the family receives psychoeducation about the disorder and its treatment, and the methylphenidate is started.

In the second and third assessment points, all instruments will be applied in screening are reapplied, except K-Sads-PL, sociodemographic questionnaire, standardized psychiatric clinic interview with standard record and NEUPSILIN-inf. In these two timepoints, we will use standardized follow-up anamnesis. Participants will also be clinically evaluated to verify the effect of the treatment (the Clinical Global Impressions (CGI) scale) [83, 84, 85], dose adjustment, the presence of adverse effects, adherence to treatment, clarification of doubts regarding the research, the disorder, and the treatment and reinforce the guidelines regarding the next steps of the study.

### Follow-up visits {11a, 18a}

Participants will also be clinically evaluated at 4, 8 and 18 weeks to verify the effect of the treatment, dose adjustment, the presence of adverse effects, adherence to treatment, clarification of doubts regarding the research, the disorder, the treatment and reinforce the guidelines regarding the next steps of the study.

The number of visits could vary, depending on the patient’s individual needs.

### Pharmacotherapy {11b}

Pharmacotherapy will be conducted by experienced child and adolescent psychiatry with primary responsibility for childcare.

Pharmacotherapy starts with a first-line stimulant medication methylphenidate. [86]. The initial dose will be prescribed as follows: 5mg in the morning and after lunch in the first week, with adjustment to 10mg at the same time until the next evaluation at 4 weeks. From that moment on, the dose will be titrated to optimize the desired therapeutic effect and minimize undesirable adverse effects. The average dose will be 1 mg/kg/day.

If the child does not tolerate the methylphenidate [87, 88] or does not respond to medication, lisdexamfetamine will be offered. [88] Other formulations may be used such as methylphenidate hydrochloride extended-release capsules or OROS methylphenidate if necessary. In these cases, equivalent doses will be prescribed.

### Blood sample {11a}

A trained nursing professional will collect three samples of venous blood from each patient/volunteer: before medication treatment, 12 and 24 weeks after starting treatment. Blood samples will be kept at a suitable temperature [89] until they are sent to the Biochemistry laboratory of the Department of Biochemistry (UFV). The BDNF [90] and the cytokines levels [91, 92, 93, 94] will be evaluated in blood plasma and the oxidative stress [95, 96] level will be evaluated in serum. Telomeres measurements will be made by DNA extraction [27, 28, 29].

### Adherence to intervention protocols {11c, 18b}

To improve adherence to medical visits and research evaluations, nursing technique will confirm the consultation by telephone (calls or WhatsApp messages) the day before. In case of participant absence, contact by phone will be made to offer appointment rescheduling.

The assessment methylphenidate adherence will be by pill count and the Treatment Adherence Measure (MAT) questionnaire [65] in all scheduled visits [97].

### Modifications {11b}

The participant will be excluded from the survey if he does not attend the assessment points (blood collection times). If the patient reports the impossibility of attending the assessment visit and is available to reschedule it within a maximum of 15 days, the patient can proceed with the intervention. The patient may miss the visits number 4, 8, 18 and proceed with the intervention, if the regular use of medication continues.

The child will be excluded in case of not tolerating the medication, family withdraws from participation, diagnosis change during follow-up.

The pharmacological intervention may have adverse effects and the child will receive all the necessary and standard care following strictly the clinical protocol. Most adverse effects are usually mild and occur early in treatment [87]. They will be informed and medication will be adjusted accordingly the need.

Participants whose diagnosis on screening differs from the diagnosis of the psychiatry team will be excluded from the study.

### Concomitant care {11d}

The treatment can also include speech therapy and occupational therapy if there is a clinical indication. The psych treatment was not available.

### Outcomes {12}

#### Primary outcomes

The BDNF levels [37], cytokines and level of oxidative stress (oxidative and antioxidant substances) after treatment with methylphenidate.

#### Secondary outcomes

Behavioral symptoms of Attention Deficit/Hyperactivity Disorder and/or Oppositional Defiant Disorder, emotional regulation level determined by the Multimodal Treatment Assessment Study - Swanson, Nolan e Pelham (MTA-SNAP-IV) scale, which assesses symptoms of attention deficit/hyperactivity disorder and Oppositional Defiant Disorder in children and adolescents, and ETDAH - parents, which assesses emotional regulation.

Cardiovascular parameters on physical examination, as well as body mass index will be recorded.

### Retention {18b}

The intervention has the potential to bring great benefits to the child and whole family. The stimulant treatment in ADHD children is associated with reduced morbidity and mortality.

WhatsApp contact will be possible for participants to clarify their doubts, reschedule the appointment, as well as be reminded of the appointments during the period in which they participate in the study.

Moreover, a report containing the child’s neuropsychological assessment will be given to the parents. In addition, all children will be under medical supervision at the outpatient clinic, even after the end of the study (UFV).

Post-intervention measures will not be necessary after the twenty-fourth week. The analysis will be performed with the information obtained up to the time of dropping out in case of child drops out of the study.

### Data Management {19}

A part of the data will be entered electronically and registered in REDCap database. A random sampling checking will be done to quality control.

Participant research files are attached to their medical records, so they will be kept for 20 years after the last registration.

### Statistical Methods {20}

#### Outcomes {20a}

The main analysis strategies will involve group comparison from the three timepoints and trajectory of symptoms and measures across time. Generalized Estimating Equations (GEE) analyses may be done with risk factors and potential moderators such social deprivation, parental styles, and oppositional symptoms to understand the relationship.

#### Additional analyses {20b}

The main additional analyses strategies will involve descriptive data to characterize the studied sample and to observe the correlations between features.

#### Analysis Population and Missing Data {20c}

Individuals that will not fill the information of the primary outcomes will have the other data analyzed: sociodemographic profile, BDNF dosages, cytokines, and oxidative stress at baseline to understand potential populational bias.

Missing data will be handled as necessary for the chosen tests, for GEE for example we will include only individuals with the complete data set for primary outcomes. We will try to retrieve all information by WhatsApp contact.

## Methods: Monitoring

### Data Monitoring {21}

#### Formal Committee {21a}

Diagnosis data was done by thirteen years experienced child psychiatry and there is a team on UFMG to discuss any doubt prof. DMM and AASJ. They also have 15 years of experience in the field.

#### Interim Analyses {21b}

Individuals that will not fill the information of the primary outcomes will have the other data analyzed: sociodemographic profile, BDNF dosages, cytokines, and oxidative stress at baseline.

Socioeconomic data will be analyzed to evaluate any potential of sampling bias.

The clinical psychiatric assessment will be done by an experienced child and adolescent psychiatry, and any diagnosis divergence will be informed. Adherence will be considered poor if the child takes less than 2/3 of prescribed doses of methylphenidate.

### Harms {22}

Adverse events during the trial will be registered in the child’s medical record and communicated to the relevant governmental agencies if necessary.

Assistance in case of adverse events will be guaranteed to the child via the public health system.

### Auditing {23}

All data will be available to audit if necessary. The diagnosis divergence will be evaluated and informed as soon as it is verified.

## Ethics and Dissemination

### Research Ethics Approval {24}

Approved by the Research Ethics Committee of the Federal University of Minas Gerais. Number: 4.364.744. CAAE: 82870117.0.3001.5149. Written, informed consent to participate will be obtained from all parents, as well as informed assent from the children.

### Protocol Amendments {25}

Any modifications to the protocol which may impact on the conduct of the study, potential benefit of the patient or may affect patient safety, will lead to a formal amendment to the protocol with REBEC, the ethics committee and clinical trial publication journal.

### Consent or Assent {26}

#### Consent or Assent {26a}

All parents must sign the free and informed consent form, as well as the children the assent form. The terms will be delivered at the end of the child’s screening if the child is considered eligible to be included in the study and they wish to participate voluntarily in the research.

#### Ancillary Studies {26b}

Not Applicable

### Confidentiality {27}

All study-related information will be stored securely at the child’s medical record. All local databases will be secured with password-protected access systems.

### Access to Data {29}

All researchers will have access to the final trial dataset without any contractual limitations.

### Ancillary and Post-trial Care {30}

The children will continue clinical treatment at the child psychiatry outpatient clinic for as long as the family wishes, or they will be discharged or reach the age of 18, when they will be referred to adult services.

### Dissemination Policy {31}

#### Trial Results {31a}

All research data and personal information will be under responsibility of the researchers to protect confidentiality before, during and after the trial. All parents or guardians’ results will be communicated at the end and regarding the trial results.

Trial results will be published at REBEC, regardless of the magnitude or direction of effect. The results will also be reported in an original article and submitted for a relevant journal.

#### Authorship {31b}

ML is the Chief Investigator; she conceived the study, led the proposal and protocol development. L, B and J contributed to development of the proposal and methodology. SC e DMM contributed to study design and to development of the proposal and methodology. All authors will read and approve the final manuscript.

#### Reproducible Research {31c}

The anonymous data information might be available under request and reasonable demand.

## Discussion

Inflammatory, neurotrophic, and oxidative parameters have been associated with the pathophysiology of ADHD. Limitations in the studies varies from small and heterogeneous samples of participants [12], lack of control for variables that may interfere with the results, such as diet, body mass index, short follow-up time, and level of physical activity, are some of these limitations. Moreover, the results are contradictory [98]. Here we propose a protocol trying to make clear clinical and biomarkers in response to medication in a cohort well characterized in a prospective follow up.

Many studies were cross-sectional and retrospective, which allows inferring only an association between inflammation and the disorder and not a causal relationship of pathogenesis [98]. Longitudinal studies are necessary to better establish the possible relationship between ADHD, inflammation, neurotrophic, and oxidative stress [16]. Other important factors to be considered are comorbid mental disorders and potential confounding factors, in addition to being important to observe possible changes in the inflammatory profile after interventions [16].

This trial protocol methodology will follow the participants by 24 weeks, increasing the chance of evaluating the influence of the chronic use of methylphenidate on inflammatory, neurotrophic, and oxidative factors. In addition, there will be evaluation of the child’s diet and physical activity through the food recall and the usual physical activity questionnaire, respectively. To reduce hormonal influences of puberty, we will exclude girls who had menarche and limited the age of both sexes to 14 years.

The results may provide information about the pathophysiology of ADHD, being able to collaborate in the identification of biomarkers of the disorder and response to treatment with methylphenidate.

## Appendices

### Appendix 1. Informed consent materials {32}

### Appendix 2. Consent and Assent forms {32}

### Appendix 3. Biorepository constitution {32}

### Biological specimens {33}

Collection and analysis of biological samples will follow standard protocols for these procedures in accordance with health surveillance recommendations.

Biorepository term was signed and contains the norms of storage and use in auxiliary and future studies. (Appendix 3)

## Data Availability

No datasets were generated or analysed during the current study. All relevant data from this study will be made available upon study completion.

## Acknowledgements

Acknowledgment to the entire Specialized Health Care team (UAES/UFV), Biochemistry and Medicine laboratory (UFV) team, Molecular Medicine laboratory (UFMG) team, scientific initiation students (UFV), psychology students (volunteer member/Univiçosa), nutrition professor and student (UFV), the neuropsychologist Roselaine da Cunha Barros (volunteer member), and the nursing techniques Arieta de Jesus Felisberto Oliveira (UFV) e Ana Cristine Pepe Parabocz (UFV).

## Supporting Information

S1_File. SPIRIT Checklist.

